# Implementation of Individualised Polygenic Risk Score Analysis: A Test Case of a Family of Four

**DOI:** 10.1101/2021.06.29.21259713

**Authors:** Manuel Corpas, Karyn Megy, Antonio Metastasio, Edmund Lehmann

## Abstract

Polygenic risk scores (PRS) have been widely applied in research studies, showing how population groups can be stratified into risk categories for many common conditions. As healthcare systems consider applying PRS to keep their populations healthy, little work has been carried out demonstrating their implementation at an individual level. We performed a systematic curation of PRS sources from established data repositories, selecting 27 phenotypes, comprising almost 40 million SNPs related to cancer, cardiovascular, metabolic and autoimmune diseases. We tested selected phenotypes using whole genome sequencing data for a family of four family related individuals, with the 1000 Genomes Project (1000G) Phase III participants as background populations. Over 98 billion allele effects were calculated in order to obtain the PRS for each of the individuals analysed here. PRS calculation for the 1000G cohort of 2,504 participants allows us to develop a methodology for risk inference and general PRS deployment. Our approach for PRS implementation advances the discussion on the adoption of PRS in a preventative healthcare setting.

## Introduction

Although genetics plays a substantial role in the development of common diseases, to date, optimising its contribution to disease prevention in individuals remains a challenge [1]. PRS are an emerging tool in genetics, the potential of which has been picked up by health systems, including in UK’s National Health Service [2], as a tool for improving the health of their populations. For some common diseases, such as Coronary Artery Disease, Type 2 Diabetes or Breast Cancer, PRS have been shown to help capture a sizable genetic contribution as part of the aetiology of high-risk individuals [3]. However, it remains to be shown how PRS can be a useful tool for disease prevention at the level of the individual in many complex conditions [4].

There have already been attempts to implement PRS in a preventative healthcare setting. For instance, the MedSeq project [5] provided a benchmark study for application of cardiovascular disease PRS in a cohort of 100 individual whole genomes. A number of direct-to-consumer companies are also providing PRS in a preventative context, including testing of traits such as Breast Cancer and Type 2 Diabetes. Nonetheless, for many of these PRS tests, only a relatively small proportion of known variants are being tested (e.g., tens or dozens), compared to the total number included in some PRS, which for Type 2 Diabetes, for instance, is in the order of 7 million Single Nucleotide Polymorphisms (SNPs) [3]. The current provision of PRS for disease risk prevention is thus not yet at the same level as in PRS research, where there is a plethora of new PRS incorporated into centralised repositories. Repositories such as Cancer-PRSweb [6] displays 69 PRS for cancer alone, while the Polygenic Score Catalog [7] reports 751 (last accessed on 24 March 2021).

Here we propose a novel implementation for reuse and deployment of PRS collected from public repositories and supported by scientific literature. Due to the heterogeneity and overlap of available PRS, we perform a systematic curation of existing data sources following a set of purposely generated criteria for their selection. We include PRS from a wide range of common diseases related to cancer, cardiovascular, metabolic and autoimmune diseases. We apply selected PRS as proof-of-principle implementation to a family of four of Iberian Spanish origin, who underwent whole genome sequencing. While we use the dataset of a family as our test case so as to be able to compare results of several related family members, we believe our methodology could be applied to a single individual.

Since we sequence the whole genome of each family member, we do not impute alleles for any variant. Instead we extract the exact allele from processed sequencing data. By using 1000 Genomes Project (1000G) individual variant data [8] as PRS background distributions, we are able to assess the genetic risk of each family member by comparing the individual’s score against the scores of the 1000G cohort.

From a total 43 PRS initially selected as candidates, we apply 27, encompassing a total of 39,126,746 tested SNPs for each family member. For each individual PRS, risk percentiles are calculated using the PRS of participants within three 1000G cohorts: Iberian Spanish (IBS; n=107), European (EUR; n=503), and all 1000G individuals (ALL; n=2,504). Over 98 billion allele effect calculations were performed in order to obtain the PRS for each of the participants used in this study. We investigate how the selection of the background 1000G PRS distributions influences tested individuals’ PRS percentile values. This allows us to identify if an individual is at the higher risk end tail of the PRS 1000G background distribution and estimate their relative risk for developing a disease.

## Methods

### Sequencing and data processing

Saliva samples were collected using Oragene OG-600 and sent for DNA extraction and sequencing. The DNA samples were randomly fragmented by Covaris technology and fragments of 350 bps were obtained. Fragment DNA ends were repaired and an ‘A’ base added at the 3’ end of each strand. Adapters were then ligated to both strands of the end repaired/dA tailed DNA fragment. Amplification by ligation-mediated PCR was performed and then single strand separation and cyclisation. DNA nanoballs were created and loaded into the patterned nanoarrays and pair-end reads read through on the BGISEQ-500 platform for each library to maximise the chances of a target of 30x coverage. Software for base calling with default parameters and the sequence data of each individual were generated as paired-end reads, identified as ‘raw data’ and provided as FASTQ format.

Once fastq files were obtained, we used the Sentieon DNASeq pipeline [9] for all four samples. Sentieon is a toolkit analogous to GATK [10] but built on a highly optimised backend. It takes raw fastq files and maps them to the human reference genome using BWA-MEM [11]. As all the PRS we were analysing used GRCh37, so we mapped to that reference. For variant calling, Sentieon uses the recommended best practices for variant analysis with GATK, with local realignment around indels and base recalibration using GATK and duplicate reads removed by Picard tools. The sequencing depth and coverage for each individual were calculated with strict data quality controls throughout the pipeline.

### Family dataset

We selected this particular family dataset because it has been well studied in the past [12–15], which affords us a deep knowledge of the family’s phenotypes and disease history. Figure 1 shows the family pedigree. In it we have individuals PT00007A (Father), PT00008A (Mother) and two children (PT00009A and PT00002A; Daughter and Son). From here onwards, and for simplicity, we refer to family members as (Father, Mother, Daughter, Son).

**Figure 1:**
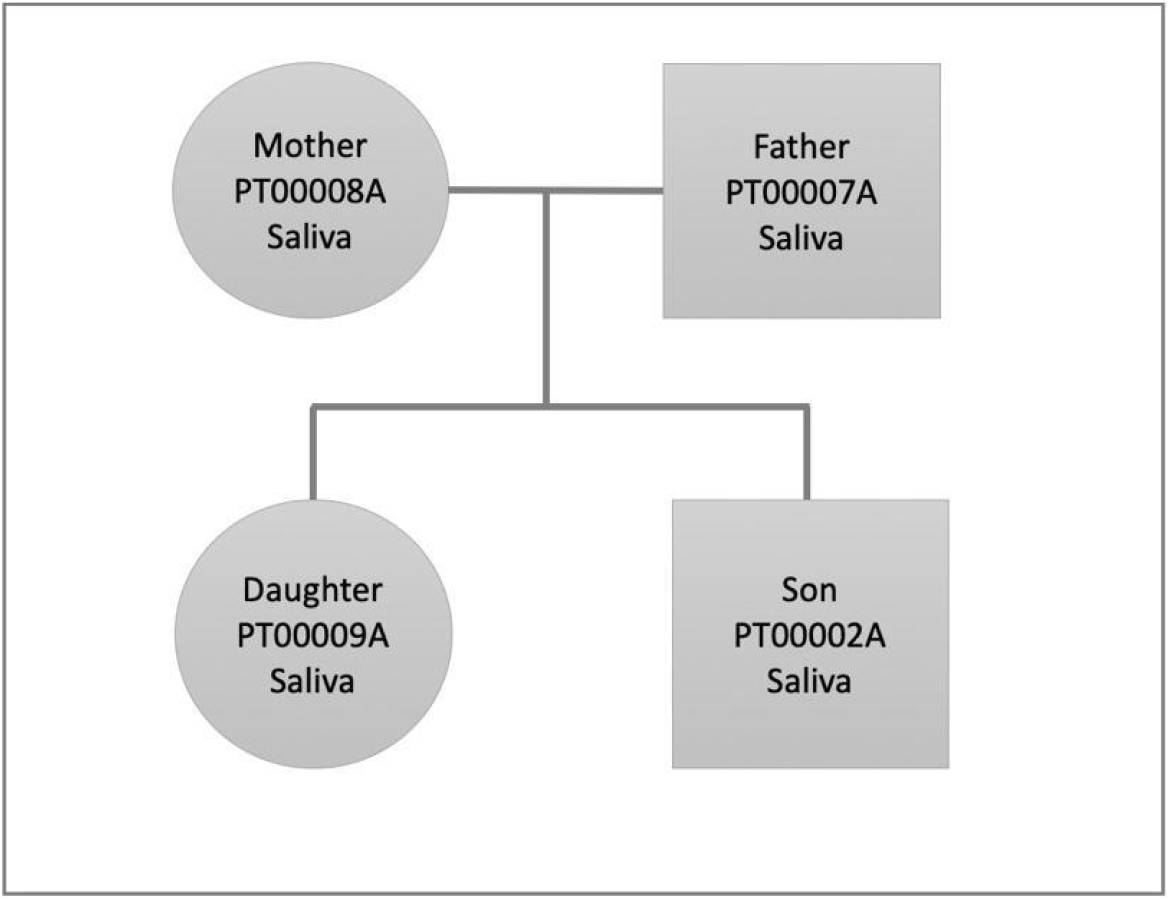
Family pedigree showing the relationship, gender (square: male, circle: female), and sample used for whole genome sequencing (saliva).

When we analysed the variant output of all samples, we benchmarked against Fabric Genomics Clinical Grade Scoring Rules (http://help.fabricgenomics.com/hc/en-us/articles/206433937-Appendix-4-Clinical-Grade-Scoring-Rules; accessed 7 / January / 2020), where Clinical Grade is a measure of a variant file’s overall quality and fitness for clinical interpretation (Table 1). Coverage in values with a star indicates that the median coverage of coding variants exceeds 40. Genotype quality with a starred value: more than 95% of the coding variants have a quality above 40. Starred homozygous / heterozygous ratio: the ratio for the coding variants is between 0.5 and 0.61. Starred transition / transversion ratio: The ratio for the coding variants is between 2.71 and 3.08. We performed a further analysis of quality of variants by counting those that pass the default standard filters of quality for interpretation given our analysis software.

**Table 1:** Statistics for clinical grade measures of the quality of the variant file. Star-marked values (*) indicates the quality is of clinical standards and no-star values that it is below clinical standards (see Fabric Genomics Clinical Grade Scoring Rules [http://help.fabricgenomics.com/hc/en-us/articles/206433937-Appendix-4-Clinical-Grade-Scoring-Rules]). The total number of variants for all saliva samples and the total number of coding variants for each family member are also shown.

| Sample ID | Coverage | Genotype Quality | Homozygous / Heterozygous ratio | Transition / Transversion Ratio | Total number of variants | Total number of coding variants |
| --- | --- | --- | --- | --- | --- | --- |
| PT00007A (Father) | 25 | 95.9* | 0.51* | 2.81* | 4,650,536 | 27,504 |
| PT00008A (Mother) | 24 | 95.7* | 0.51* | 2.79* | 4,695,886 | 27,329 |
| PT00009A (Daughter) | 29 | 97.8* | 0.48 | 2.79* | 4,812,818 | 27,400 |
| PT00002A (Son) | 43.0* | 94.3 | 0.51* | 2.81* | 4,956,742 | 27,286 |

### Ethical framework

All participants underwent a consent process and signed a consent form accepting the terms and conditions of this work as well as the potential consequences of performing this analysis. We drew on the Personal Genome Project UK [16] as our approach to informed consent. The consent process we developed included the following elements: (a) participants underwent extensive training on the risks of genetic analysis including the risks of publishing personal genetic data; (b) participants completed an exam to demonstrate their comprehension of the risks and protocols associated with participating in genetic analysis which may be published and (c) participants were judged truly capable of giving informed consent. Consent forms were signed by all family members. This ethical framework has been independently assessed and approved by the Ethics Committee of Universidad Internacional de La Rioja (code PI:029/2020).

### Curation of PRS

Underlying PRS data have been made available by the scientific community through the Polygenic Score Catalog [7] and Cancer-PRSweb [6]. Both resources provide centralised access to many PRS as well as data needed for their application, including SNP coordinates, effect alleles and their effect weights. We performed a curation process to identify PRS for application to our family use case of four. A dataset of 39,126,746 PRS SNPs was generated encompassing 27 common diseases (we call these common diseases ‘phenotypes’ from now onwards), together with risk alleles and weighted contributions for each SNP. Table 2 shows the two sets of criteria we followed to select PRS for our implementation model. The first set of criteria is based on study design and performance (Table 2a: Design Selection Criteria) and second on the requirements needed for their bioinformatics implementation (Table 2b: Bioinformatics Selection Criteria). In terms of design criteria, we chose PRS whose characteristics matched the following properties: i) Underlying GWAS: the Genome Wide Association Study (GWAS) underlying the PRS could be traced to a recognisable consortium, and the phenotype in the GWAS was consistent with the phenotype in the resulting PRS. ii) The PRS was trained in a second study using previously published PRS creation methods (e.g., LDpred [17] or clumping and thresholding [18]). iii) The PRS was validated in a large independent cohort (we developed a preference for the UK Biobank for consistency reasons) [19]. Its area under the curve (AUC) or similar performance metric is above the 0.55 threshold. v) In case of more than one PRS being available for the same phenotype, we made a judgement of the study as a whole. vi) The PRS should ideally have published risk metrics such as odds ratios, hazard ratios or fold increase.

**Table 2.** Criteria we used to select Polygenic Risk Scores (PRS) for our study. GWAS: Genome Wide Association Study; AUC: Area Under the Curve; 1000G: 1000 Genomes Project; hg19: Human Genome 19 reference assembly.

| 2a. PRS Design Selection Criteria |
| --- |
| i. Traceable to a discovery GWAS from a recognised consortium |
| ii. Trained in a second study using previously published PRS creation methods |
| iii. Validation in a large independent cohort; preferably UKB |
| iv. AUC or similar performance metric above 0.55 |
| v. If several PRS of same phenotype: judgement of study as a whole |
| vi. Have risk metrics if possible: odds ratios, hazard ratios, fold increase |

| 2b. Bioinformatic Selection Criteria |
| --- |
| i. At least 95% SNPs present in 1000G Phase III distribution |
| ii. Pass matching allele filter |
| iii. Have effect weights |
| iv. Coordinates in hg19 |

Once we had filtered out PRS that did not fulfil the above design criteria, the remaining phenotypes underwent an extra filtering process according to a set of bioinformatics standards (Table 2b), required for us to run our pipelines successfully. These bioinformatics filtering criteria involved processing of the PRS raw data to establish that they fulfilled the following conditions: i) Presence of at least 95% of SNPs in the 1000G Phase III distribution. ii) Presence of risk alleles in either the reference or the alternative allele of the 1000G matching variant annotation. For this we check whether each SNP risk allele has an exact match to the reference or alternative allele in that coordinate position, discarding and labelling the SNP as ‘missing’ if otherwise (this enabled us to identify any reverse strand or similar bioinformatic inconsistencies) iii) availability of effect weights and iv) availability of coordinates in hg19. We used hg19 coordinates due to PRS source data being made available using this genome assembly. Any SNP that did not meet the above criteria was discarded but the phenotype was still used as long as it retains at least 95% of its source PRS SNPs.

### How we calculate PRS for an individual

Our first step in calculating a PRS for a family member was to create background distributions so as to be able to put the score of a family member into context, and thus understand his or her relative risk. This is because source publications do not offer a translation of a raw PRS score directly into a risk measurement. Rather, they stratify different sections of a studied group into risk buckets (for example the top 5% of a distribution may be ascribed a particular odds ratio (OR)). Hence, when applying a PRS to an individual, it is necessary to know where that individual sits relative to others.

A PRS was calculated for each individual as the sum of the effect weights for all the risk alleles observed in the individual for a particular phenotype, divided by the total number of risk alleles reported for that phenotype. We calculated PRS following this method for each of the individuals in the final (Phase III) dataset of the 1000 Genomes Project (1000G), containing data for 2,504 participants. This required us to calculate the PRS of all 2,504 individuals in the 1000G project across all selected phenotypes.

Having generated raw scores for each of these 1000G individuals, we built distributions of raw scores according to different population groups within the 1000G cohort. We chose the subset of Iberians Spanish (IBS, n=107), as all family members are of Spanish origin, the Europeans (EUR, n=503), reflecting the ethnic background of the validation data sets for the PRS we selected, and the entire 1000G cohort (ALL, n=2,504). ALL contains African, Admixed American, East Asian, South Asian and Europeans.

We then applied the same methodology for calculating a raw PRS score to the whole genome data of each of the four family members, and having determined the raw score for each family member for each phenotype, we placed that score inside the distribution of each population group already generated from the 1000G individuals (IBS, EUR and ALL).

Placing the individual in context this way allowed us to derive percentiles reflecting a family member’s position in a given population for a given phenotype. We could then readily compare these percentiles between individuals for each phenotype. We did this across the three different population groups in order to control for the impact of the ethnicity of the background population on the resulting percentile.

Scores from both 1000G participants and family members are thus calculated independently, producing a distribution of scores from which the percentile a family member occupies is generated.

### PRS percentile inheritance patterns

We were interested to understand patterns of inheritance among family individuals for PRS percentiles. We set out to analyse how a high or low PRS percentile was explained in terms of risk being passed on from parents to children. This is a useful quality control and a way of adding credence to results, since it would be strange if for the same phenotype low risk observed in both parents engender high risk in offspring. In order to compare PRS percentiles between family individuals, we compare values relative to the 1000G EUR population distribution. We choose the EUR distribution percentile values for the remaining analyses because all selected PRS have as their validation dataset a European population such as those contained in the UK Biobank or FINRISK [20].

For the purposes of this analysis only, we have defined high risk as the individual falling above the 80th percentile of a given background distribution, as this is the threshold from which both Khera et al. [3] and Mars et al. [21] begin to quantify elevated risk. However, we acknowledge that the definition of a high-risk percentile is somewhat subjective, and we expand on this further below.

### Translation of percentiles into risk metrics

For each family member we ascribe a relative risk. We note that when translating PRS percentiles into genetic risk metrics, each phenotype must be interpreted differently, as the risk metrics (e.g., odds ratios or hazard ratios), and risk thresholds vary from study to study. If a family member’s percentile is within a reported threshold of the PRS percentile source publication, we attribute a risk metric to that family member (Table 7). We also pay attention to the Area Under the Curve (AUC) or other performance metrics described by each PRS source study.

### Impact of population background distributions on risk percentiles

We considered the effect of background populations in risk calculation. There is a known risk that PRS predict less well in populations where the underlying GWAS and validation cohorts differ from the ancestry of the individual [22], as SNPs have different allele frequencies depending on ancestry. Therefore, an individual may be assigned different percentiles depending on background populations. This is important, given that odds ratios or hazard ratios are reported relative to intervals in PRS percentiles. If the choice of background population significantly changes an individual’s percentile PRS (i.e., >20 percentile), their resulting odds or hazard ratio will then be different, affecting how we interpret risk. In order to determine whether or not the choice of background population made a difference to the results, we performed two types of analyses:

1. **Analysis of phenotype percentiles individually**. We checked whether there are any noticeable differences in individual phenotype PRS percentiles depending on the choice of background distribution for each family member. For this, we compare whether tested individual percentiles for a phenotype change PRS quintiles depending on their background distribution. This choice of quintiles for binning risk distributions is a popular thresholding among the studies we curated [3,21].
2. **Statistical correlations between populations across all phenotypes**. We tested the strength of the differences between any pair of distributions (IBS vs EUR, IBS vs ALL and EUR vs ALL) for each family individual via Pearson correlation coefficients. This analysis was done across the whole group of 27 phenotypes, rather than phenotype by phenotype.

## Results

### PRS Curation

Our first set of criteria for selection of PRS considered the characteristics of the source study design, including recognisable GWAS consortia, performance metrics, presence of risk boundaries and independent cohort validation. We did not curate every single phenotype available, only those we judged promising candidates. Table 3 includes all phenotypes we researched after initial shortlisting. From an initial list of 43, we discarded 13 because a) they was not a clear consistency between the phenotype of the PRS and the phenotype in the underlying GWAS (for example All Cause Mortality, where the PRS is a composite of many separate GWAS); b) there was an alternative better performing candidate for the same phenotype (e.g., Coronary Artery Disease, Breast Cancer or Prostate Cancer); c) their performance metrics were below our acceptable threshold (e.g., Epithelial Ovary Cancer, Cancer of other Lymphoid, Histiocytic Tissue, Cancer of Kidney); d) their validation population was not the UK Biobank. We began with the PGS Catalog, and then complemented our selected set of PRS with Cancer-PRSweb phenotypes. From the Cancer-PRSweb we only considered their top 20 UK Biobank validated PRS, comparing them with phenotypes in the PGS Catalog where we found overlap. We tended to favour selection of standardised PRS such as those offered by the larger studies or the Cancer-PRSweb, as its blocks of performance metrics, risk boundaries, percentile thresholds and validation cohort metadata are well suited for benchmarking.

**Table 3:**
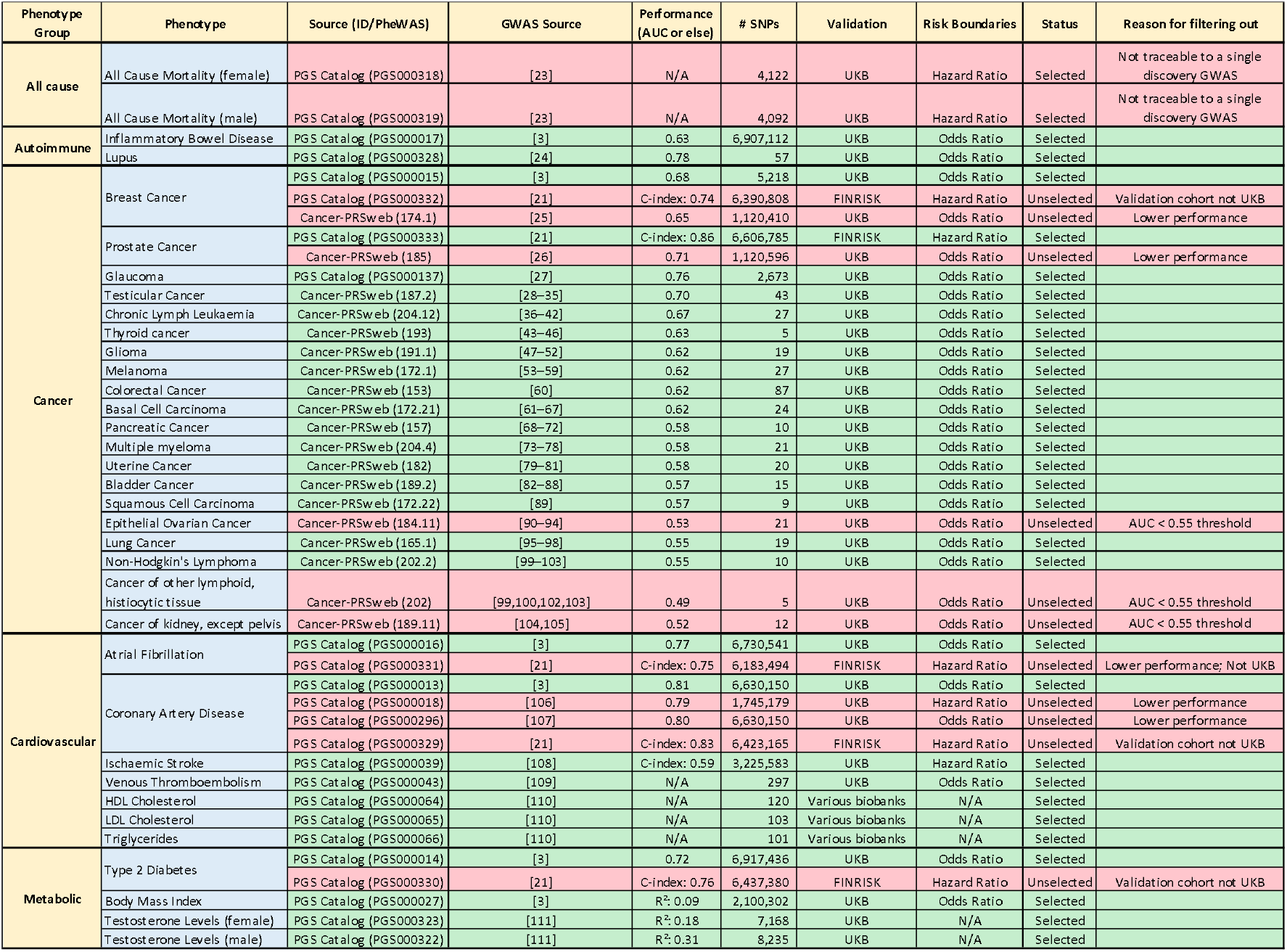
Initial list of PRS. Phenotypes are grouped according to the type of disease they relate to (e.g., all-cause, autoimmune, cancer, cardiovascular and metabolic), the source Genome Wide Association Study (GWAS) Consortium, performance metrics (AUC or an alternative if possible), number of total SNPs, the cohort used for their validation (UKB: UK Biobank), reported risk metric and the reason for filtering them out if unselected.

We had to reconcile conflicting criteria in the cases of Breast Cancer and Prostate Cancer PRS selection, and here we did deploy our judgement. For Breast Cancer, we selected the PRS from Khera et al. [3], although it has a lower performance than Mars et al. [21]. This was because overall the PRS for Khera et al. are high performing, and are all validated in the UK Biobank. For consistency therefore we retained the Breast Cancer phenotype from the Khera et al. study. For Prostate Cancer, despite being validated in the FINRISK consortium, we decided that the C-index of 0.86 in the Mars et al. study was sufficiently differentiated against that of Cancer-PRSweb (AUC of 0.71) that the Mars et al. PRS merited selection.

Concerning AUCs, we allowed any covariates that the source GWAS studies allowed. We recognise that this means that an AUCs in one PRS is not exactly comparable to an AUC for another, as their design is not identical. Furthermore, we do not make a distinction between the type of method applied to calculation of the PRS (e.g., LDPred, Pruning and thresholding, etc.), accepting any method as long as it has been peer reviewed. Finally, we also note that some phenotypes are discrete while others are not (e.g., BMI), further affecting the choice of PRS calculation method.

Having made an initial selection of phenotypes whose study design met our eligibility criteria, we applied Table 2b’s bioinformatic filtering criteria scheme (Table 4). These bioinformatic requirements derived from the need to reliably replicate a PRS percentile as originally envisaged by the source publication. Because we use the 1000 Genomes (1000G) Project Phase III participants as our background PRS distributions, we required a high overlap (>95%) of all PRS effect alleles and their weights between the SNPs identified by the study in question and the 1000G project. We also limited our selection PRS to only those that use autosomal SNPs.

**Table 4:** Our set of bioinformatic filtering criteria applied to the remaining phenotypes.

| Phenotype Group | Phenotype (PheWAS Code) | Source (ID/PheWAS) | GWAS Consortium | # SNPs | Missing SNPs | % Missing SNPs | Validation | Status | Reason for filtering out |
| --- | --- | --- | --- | --- | --- | --- | --- | --- | --- |
| Autoimmune | Inflammatory Bowel Disease | PGS Catalog (PGS000017) | [3] | 6,907,112 | - | - | UKB | Selected |  |
|  | Lupus | PGS Catalog (PGS000328) | [24] | 57 | 32 | 56.14% | UKB | Unselected | Missing SNPs > 5% |
| Cancer | Breast Cancer | PGS Catalog (PGS000015) | [3] | 5,218 | - | - | UKB | Selected |  |
|  | Prostate Cancer | PGS Catalog (PGS000333) | [21] | 6,606,785 | 832 | 0.01% | FINRISK | Selected |  |
|  | Glaucoma | PGS Catalog (PGS000137) | [27] | 2,673 | 16 | 0.60% | UKB | Selected |  |
|  | Testicular Cancer | Cancer-PRSweb (187.2) | [28–35] | 43 | - | - | UKB | Selected |  |
|  | Chronic Lymph Leukemia | Cancer-PRSweb (204.12) | [36–42] | 27 | - | - | UKB | Selected |  |
|  | Thyroid cancer | Cancer-PRSweb (193) | [43–46] | 5 | - | - | UKB | Selected |  |
|  | Glioma | Cancer-PRSweb (191.1) | [47–52] | 19 | - | - | UKB | Selected |  |
|  | Melanoma | Cancer-PRSweb (172.1) | [53–59] | 27 | 1 | 3.70% | UKB | Selected |  |
|  | Colorectal Cancer | Cancer-PRSweb (153) | [60] | 87 | 1 | 1.15% | UKB | Selected |  |
|  | Basal Cell Carcinoma | Cancer-PRSweb (172.21) | [61–67] | 24 | 1 | 4.17% | UKB | Selected |  |
|  | Pancreatic Cancer | Cancer-PRSweb (157) | [68–72] | 10 | - | - | UKB | Selected |  |
|  | Multiple myeloma | Cancer-PRSweb (204.4) | [73–78] | 21 | - | - | UKB | Selected |  |
|  | Uterine Cancer | Cancer-PRSweb (182) | [79–81] | 20 | - | - | UKB | Selected |  |
|  | Bladder Cancer | Cancer-PRSweb (189.2) | [82–88] | 15 | - | - | UKB | Selected |  |
|  | Squamous Cell Carcinoma | Cancer-PRSweb (172.22) | [89] | 9 | - | - | UKB | Selected |  |
|  | Lung Cancer | Cancer-PRSweb (165.1) | [95–98] | 19 | - | - | UKB | Selected |  |
|  | Non-Hodgkin's Lymphoma | Cancer-PRSweb (202.2) | [99–103] | 10 | - | - | UKB | Selected |  |
| Cardiovascular | Atrial Fibrillation | PGS Catalog (PGS000016) | [3] | 6,730,541 | - | - | UKB | Selected |  |
|  | Coronary Artery Disease | PGS Catalog (PGS000013) | [3] | 6,630,150 | - | - | UKB | Selected |  |
|  | Ischaemic Stroke | PGS Catalog (PGS000039) | [108] | 3,225,583 | 11,103 | 0.34% | UKB | Selected |  |
|  | Venous Thromboembolism | PGS Catalog (PGS000043) | [109] | 297 | - | - | UKB | Selected |  |
|  | HDL Cholesterol | PGS Catalog (PGS000064) | [110] | 120 | - | - | Various biobanks | Selected |  |
|  | LDL Cholesterol | PGS Catalog (PGS000065) | [110] | 103 | - | - | Various biobanks | Selected |  |
| Metabolic | Triglycerides | PGS Catalog (PGS000066) | [110] | 101 | - | - | Various biobanks | Selected |  |
|  | Type 2 Diabetes | PGS Catalog (PGS000014) | [3] | 6,917,436 | - | - | UKB | Selected |  |
|  | Body Mass Index | PGS Catalog (PGS000027) | [3] | 2,100,302 | - | - | UKB | Selected |  |
|  | Testosterone Levels (female) | PGS Catalog (PGS000323) | [111] | 7,168 | - | - | UKB | Unselected | Missing SNPs > 5% |
|  | Testosterone Levels (male) | PGS Catalog (PGS000322) | [111] | 8,235 | - | - | UKB | Unselected | Missing SNPs > 5% |

A total of 27 phenotypes passed all our selection criteria for PRS implementation and testing. These phenotypes involved conditions related to cancer, cardiovascular, metabolic and autoimmune diseases. 21 of these phenotypes summed less than 10,000 (8,837) SNPs in total, whereas 7 phenotypes composed the vast majority of tested SNPs (39,117,909; 99.97%). We note that 11,954 SNPs were missing from our PRS calculation because they were not present as 1000G variants or their risk allele did not match the 1000G reference or alternative allele. However, the missing number of SNPs was never greater than 5% of the total for any of our selected phenotypes. The vast majority of PRS missed significantly fewer than 5% of the SNPs, and in fact more often than not, no SNPs were missed (21 out of 27 phenotypes missed none). The phenotype that proportionally misses the greatest number of SNPs is Basal Cell Carcinoma (missing 1 out of 24 SNPs; 4.17%), whereas Ischaemic Stroke missed the greatest absolute number: 11,103 SNPs (0.34%). We also note that (apart from the Cholesterols and Triglycerides, for which curation options were limited) all of the applied PRS were validated in the UK Biobank, with the exception of Prostate Cancer, which was validated on the FINRISK population.

### Patterns of risk inheritance among family members

When analysing patterns of inheritance, whenever we find high risk percentiles (80^th^ percentile or above) in both parents, we do not find any PRS that is not high risk in both children as well (Table 5). In addition, we do not find high risk in the children that is not present in either parent except for Thyroid Cancer in Son. For this phenotype, both parents harbour average PRS percentiles and therefore this high risk could be explained as a combination of different allele risk from both parents.

**Table 5:** Phenotype PRS percentiles for each family individual. Lighter shades of green highlight percentiles above or below 80 and 20 (respectively). Darker colour shades indicate percentiles in the top and bottom 5^th^ risk percentile. Phenotypes in the table have been ordered to highlight patterns.

| Phenotype | PT00007A (Father) | PT00008A (Mother) | PT00009A (Daughter) | PT00002A (Son) |
| --- | --- | --- | --- | --- |
| Non-Hodgkin's Lymphoma | 93.84 | 95.03 | 92.25 | 95.03 |
| Colorectal Cancer | 97.42 | 79.92 | 97.22 | 91.05 |
| Triglycerides | 91.85 | 65.21 | 83.90 | 63.22 |
| Coronary Artery Disease | 29.62 | 96.62 | 89.86 | 81.91 |
| Uterine Cancer | 71.77 | 78.93 | 82.90 | 56.66 |
| Testicular Cancer | 42.35 | 90.66 | 95.83 | 38.37 |
| Glaucoma | 88.67 | 42.35 | 65.01 | 65.21 |
| LDL Cholesterol | 80.12 | 62.43 | 94.04 | 19.68 |
| Type 2 Diabetes | 60.44 | 71.37 | 68.79 | 45.33 |
| Prostate Cancer | 50.50 | 80.91 | 92.45 | 21.67 |
| HDL Cholesterol | 24.45 | 82.11 | 42.94 | 86.08 |
| Pancreatic Cancer | 30.02 | 79.32 | 45.33 | 70.18 |
| Thyroid Cancer | 49.30 | 50.50 | 32.41 | 90.66 |
| Breast Cancer | 17.30 | 85.69 | 53.28 | 64.61 |
| BMI | 36.78 | 83.30 | 58.85 | 38.77 |
| Ischaemic Stroke | 26.24 | 82.90 | 64.02 | 38.17 |
| Inflammatory Bowel Disease | 43.54 | 70.78 | 46.72 | 46.72 |
| Venous Thromboembolism | 15.71 | 78.13 | 55.07 | 55.07 |
| Squamous Cell Carcinoma | 18.89 | 58.65 | 37.18 | 58.65 |
| Bladder Cancer | 18.29 | 35.79 | 69.58 | 45.92 |
| Chronic Lymph Leukaemia | 16.50 | 45.53 | 41.15 | 44.14 |
| Lung Cancer | 13.32 | 25.05 | 22.66 | 37.38 |
| Basal Cell Carcinoma | 55.86 | 7.55 | 10.54 | 14.12 |
| Multiple myeloma | 5.37 | 17.50 | 15.71 | 44.53 |
| Glioma | 4.57 | 42.35 | 18.69 | 5.96 |
| Melanoma | 14.51 | 9.74 | 22.86 | 23.26 |
| Atrial Fibrillation | 16.50 | 15.51 | 8.55 | 7.16 |

We observe several patterns of PRS inheritance among family members:

1. **A high-risk in both parents is inherited by both offspring**. Examples of this pattern are observed in the Colorectal Cancer and Non-Hodgkin’s Lymphoma phenotypes. Here, Father and Mother have a high risk which is passed to both children.
2. **High risk for one parent is inherited by one or both children**. Mother has a high risk of Coronary Artery Disease. Both children display a high risk as well. For Triglycerides, Father has a high risk and Mother a lower risk. The high risk is inherited by Daughter but not Son.
3. **High risk in a parent gets diluted in offspring through contribution from the other parent**. For Glaucoma, Father has a high risk and Mother an intermediate risk. Both children display an intermediate risk. Father has a low risk of Venous Thromboembolism, while Mother’s is high. Both children have intermediate risk. For Ischaemic Stroke and BMI, Father has a low risk, Mother has high and both children are predicted an intermediate risk.
4. **Low risk in both parents is inherited by both offspring**. This is the case of Atrial Fibrillation, Glioma, Lung Cancer, Melanoma, Multiple Myeloma and Pancreatic Cancer.
5. **Mild risk from parents leads to increased risk in offspring**. Neither Father nor Mother qualify as high-risk carriers for Prostate Cancer, yet Daughter (who is unable to express the phenotype) is predicted to be high-risk.

### Translation of percentiles into risk metrics

When consulting source publications to ascertain the risk of developing a disease phenotype given a particular percentile, we found that risks and their thresholds are differently described depending on the publication.

Khera et al. (2018) [3] for phenotypes Breast Cancer, Atrial Fibrillation, Coronary Artery Disease, Type 2 Diabetes and Inflammatory Bowel Disease offer odds ratios for patients in the top 20%, 10%, 5%, 1% and 0.5% of the distribution of risk versus the remaining part of the distribution (80%, 90%, 95%, 99%, 99.5%) as the reference group. 95% confidence intervals and P-values are also provided.

All Cancer-PRSweb phenotypes offer odds ratios for the top 25%, 10%, 5%, 2% and 1% of the PRS distribution versus the rest, together with 95% confidence intervals and P-values.

The source study led by Craig et al. [27], from which we take the Glaucoma phenotype provides odds ratios for the top 50%, 20%, 10%, 5%, 2% and 1% versus the rest of the distribution. This study offers odds ratios for Father, whose risk percentile is 88.675, but also for individuals above 50%, i.e., Daughter and Son’s, whose percentile risks are 65.01 and 65.21, respectively.

For the Kuchenbaecker et al. (2019) [110] phenotypes (HDL Cholesterol, LDL Cholesterol, Triglycerides), we were unable to retrieve risk ratios from the publication.

From Mars et al. (2020) [21] we selected their Prostate Cancer PRS, extracting odds ratios and 95% confidence intervals per standard deviation increase, using the FINRISK (n=21,813) population as the validation dataset. (We did not use other phenotypes from this publication as they are already covered by Khera et al. (2018)[3] and we decided to choose PRS from the latter).

Abraham et al. (2019) [108], which studies Ischaemic Stroke, does not provide odds ratios. Instead, they offer a hazard ratio per standard deviation by age 75, using the UKB as the validation dataset.

Finally, Khera et al. (2019) [112] when considering BMI, report the top 10% of the distribution of high PRS carriers for different BMI levels: ≥40, ≥50 and ≥60 kg/m2.

For each extracted odds/hazard ratio, each source publication must be considered independently when reporting for an individual. In most cases, the PRS percentile of the individual lies within a reported interval from the source thresholds, but there are exceptions. For instance, the BMI source study, which only reports for the top decile, ends up being annotated for Mother as having an odds ratio <4.22 as this value is for 90 PRS percentile or above (Mother=83.30%).

Percentile thresholds vary from ≥50% to ≥99.5%, depending on the source publication. We note all lower end confidence intervals as being >1 and P-values much lower than the significance threshold of 0.05 (risks very likely not to have occurred by chance). Table 6 summarises source publication extracted risks based on PRS percentiles for each family member.

**Table 6:** Risk ratios (Odds Ratio (OR) or Hazards Ratio (HR)) extracted from PRS sources. ORs and confidence intervals are dependent on the individual’s position in the background population and are translated into risk metrics based on boundaries of bins provided in the relevant study, rather than being a standalone assessment of the individual’s risk. Blank cells correspond to phenotypes where an individual’s EUR background population percentile is below reported thresholds in PRS sources. We highlight in red risk ratios (OR or HR) we can express and in yellow those that are either not available (N/A; high percentile but no risk provided) or cannot be expressed by the individual (e.g., Testicular Cancer and Prostate Cancer PRS in females). OR or HR appear with their 95% confidence intervals (in parenthesis) and, in a separate column, the percentile thresholds from which risk ratios were extracted.

| Phenotype | Father Risk<br>(95% CI) | Mother Risk<br>(95% CI) | Daughter Risk<br>(95% CI) | Son Risk<br>(95% CI) | Risk Type | Thresholds |
| --- | --- | --- | --- | --- | --- | --- |
| Basal Cell Carcinoma |  |  |  |  |  |  |
| Bladder Cancer |  |  |  |  |  |  |
| Breast Cancer |  | 2.07<br>(1.97-2.19) |  |  | OR | Top 20% vs Rest |
| Chronic Lymph Leukaemia |  |  |  |  |  |  |
| Colorectal Cancer | 2.69<br>(2.34-3.08) | 2.69<br>(2.34-3.08) | 2.69<br>(2.34-3.08) | 2.69<br>(2.34-3.08) | OR | Top 25% vs Rest |
| Glaucoma | 3.61<br>(3.11-4.20) |  | 2.94<br>(2.60-3.34) | 2.94<br>(2.60-3.34) | OR | Top 20% vs Rest<br>Top 50% vs Rest |
| Glioma |  |  |  |  |  |  |
| HDL Cholesterol |  | N/A |  | N/A | - |  |
| LDL Cholesterol |  |  | N/A |  | - |  |
| Lung Cancer |  |  |  |  |  |  |
| Melanoma |  |  |  |  |  |  |
| Multiple myeloma |  |  |  |  |  |  |
| Non-Hodgkin's Lymphoma | 2.05<br>(1.62-2.61) | 2.05<br>(1.62-2.61) | 2.05<br>(1.62-2.61) | 2.05<br>(1.62-2.61) | OR | Top 10% vs Rest<br>Top 5% vs Rest |
| Pancreatic Cancer |  | 1.8<br>(1.19-2.72) |  |  | OR | Top 25% vs Rest |
| Squamous Cell Carcinoma |  |  |  |  |  |  |
| Testicular Cancer |  | 3.69<br>(2.2-6.18) | 3.69<br>(2.2-6.18) |  | OR | Top 10% vs Rest |
| Thyroid Cancer |  |  |  | 3.48<br>(2.16-5.62) | OR | Top 10% vs Rest |
| Triglycerides | N/A |  | N/A |  | - |  |
| Uterine Cancer |  | 1.6<br>(1.17-2.19) | 1.6<br>(1.17-2.19) |  | OR | Top 25% vs Rest |
| Venous Thromboembolism |  |  |  |  |  |  |
| Prostate Cancer |  | 2.29<br>(1.75-3.00) | 2.29<br>(1.75-3.00) |  | OR | >1SD |
| Ischaemic Stroke |  | 1.26<br>(1.22-1.31) |  |  | HR | >1SD |
| Atrial Fibrillation |  |  |  |  |  |  |
| BMI |  | < 4.22<br>(3.98-4.49) |  |  | OR | Top 10% vs Rest<br>BMI ≥ 40 |
| Coronary Artery Disease |  | 3.34<br>(3.12-3.58) | 2.55<br>(2.43-2.67) | 2.55<br>(2.43-2.67) | OR | Top 20% vs Rest<br>Top 5% vs Rest |
| Type 2 Diabetes |  |  |  |  |  |  |
| Inflammatory Bowel Disease |  |  |  |  |  |  |

### Effect of background population in percentile calculation

In order to test whether the choice of background population for percentile calculations are significantly different, we conducted two types of analyses:

1. **Analysis of phenotype percentiles individually**. We checked whether there are any significant differences at the level of individual phenotypes when comparing the effect of background PRS distributions. Table 7 depicts a heat map highlighting phenotypes for family members (Father, Mother, Daughter, Son) in the top (red) and bottom (green) PRS quintiles using three background 1000G distributions (IBS, EUR and ALL). Stronger shades of red/green are shown for phenotype PRS in the top 5^th^ or bottom 5^th^ percentile, respectively. We observe that the pattern of red/green, although generally conserved across the three background distributions and between family members, also show differences. Differences within the same individual reflect how the PRS percentile changes when comparing it against a different 1000G population group. For example, we see differences in Basal Cell Carcinoma, BMI, Ischaemic Stroke and Type 2 Diabetes. Primarily, these differences follow two patterns: a) lower percentiles for IBS/EUR than ALL; e.g., Basal Cell Carcinoma; b) higher percentiles in IBS/EUR and lower for ALL; e.g., BMI, Ischaemic Stroke and Type 2 Diabetes, with all family individuals having much higher percentiles in the IBS and EUR background distribution than ALL. We also observe similar percentiles when comparing across distributions. We note as examples Colorectal Cancer, Coronary Artery Disease and Non-Hodgkin’s Lymphoma, where a family member’s PRS percentile is similar across the different background distributions. Whether the percentile PRS is consistent among different populations does not depend on the source study. For instance, looking at results of PRS from Khera et al. (2018) [3] Type 2 Diabetes gives inconsistent results (i.e., quintiles differing by >20 percentile points) across population groups, while Coronary Artery Disease gives greater consistency. Similarly, we find consistent percentiles in Cancer-PRSweb phenotypes (Non-Hodgkin’s Lymphoma) and inconsistent ones (Basal Cell Carcinoma).
2. **Statistical correlations between populations across all phenotypes**. We tested correlations between any pair of background distributions (IBS vs EUR, IBS vs ALL and EUR vs ALL) in each family member across all phenotypes. Table 8 shows the results of these correlation tests. We find a high correlation between IBS and EUR in all family individuals [Father (0.97), Mother (0.98), Daughter (0.98) and Son (0.97)]. However, when we compare ALL with IBS and EUR, we find lower correlations in the results of each family member. The lowest observed correlation is in Mother (EUR vs ALL; R=0.44, IBS vs ALL R=0.47).

**Table 7:** Summary of percentile PRS using IBS, EUR and ALL background distributions for each family member. We highlight the top 80^th^ PRS percentile (red) and bottom 20^th^ percentile (green). Stronger shades of red/green are depicted for phenotype PRS in the top or bottom 5^th^ percentile, respectively.

|  | PT00007A (Father) |  |  | PT00008A (Mother) |  |  | PT00009A (Daughter) |  |  | PT00002A (Son) |  |  |
| --- | --- | --- | --- | --- | --- | --- | --- | --- | --- | --- | --- | --- |
|  | IBS (n=107) | EUR (n=503) | ALL (n=2504) | IBS (n=107) | EUR (n=503) | ALL (n=2504) | IBS (n=107) | EUR (n=503) | ALL (n=2504) | IBS (n=107) | EUR (n=503) | ALL (n=2504) |
| Colorectal Cancer | 99.07 | 97.42 | 99.36 | 86.92 | 79.92 | 88.94 | 99.07 | 97.22 | 98.96 | 98.13 | 91.05 | 95.25 |
| Non-Hodgkin's Lymphoma | 92.52 | 93.84 | 90.58 | 93.46 | 95.03 | 91.09 | 90.65 | 92.25 | 88.30 | 93.46 | 95.03 | 92.17 |
| Coronary Artery Disease | 30.84 | 29.62 | 44.69 | 99.07 | 96.62 | 96.17 | 94.39 | 89.86 | 88.78 | 86.92 | 81.91 | 81.99 |
| Uterine Cancer | 76.64 | 71.77 | 67.53 | 82.24 | 78.93 | 76.16 | 85.98 | 82.90 | 80.31 | 60.75 | 56.66 | 56.27 |
| Triglycerides | 91.59 | 91.85 | 79.83 | 66.36 | 65.21 | 42.53 | 85.05 | 83.90 | 67.05 | 65.42 | 63.22 | 41.01 |
| LDL Cholesterol | 79.44 | 80.12 | 90.18 | 57.94 | 62.43 | 73.88 | 95.33 | 94.04 | 97.92 | 15.89 | 19.68 | 30.35 |
| Testicular Cancer | 37.38 | 42.35 | 44.01 | 88.79 | 90.66 | 91.17 | 96.26 | 95.83 | 95.89 | 31.78 | 38.37 | 40.65 |
| Glaucoma | 89.72 | 88.67 | 83.91 | 42.06 | 42.35 | 29.47 | 69.16 | 65.01 | 56.51 | 70.09 | 65.21 | 56.71 |
| Thyroid Cancer | 56.07 | 49.30 | 59.74 | 57.01 | 50.50 | 60.10 | 40.19 | 32.41 | 39.70 | 95.33 | 90.66 | 95.21 |
| Squamous Cell Carcinoma | 26.17 | 18.89 | 79.15 | 57.01 | 58.65 | 90.18 | 44.86 | 37.18 | 84.66 | 57.01 | 58.65 | 90.18 |
| Prostate Cancer | 51.40 | 50.50 | 40.06 | 81.31 | 80.91 | 62.86 | 91.59 | 92.45 | 72.96 | 22.43 | 21.67 | 16.57 |
| Inflammatory Bowel Disease | 63.55 | 43.54 | 37.50 | 88.79 | 70.78 | 60.86 | 66.36 | 46.72 | 40.65 | 66.36 | 46.72 | 40.65 |
| HDL Cholesterol | 17.76 | 24.45 | 15.26 | 79.44 | 82.11 | 70.81 | 36.45 | 42.94 | 31.91 | 84.11 | 86.08 | 76.08 |
| Venous Thromboembolism | 15.89 | 15.71 | 12.34 | 83.18 | 78.13 | 86.22 | 64.49 | 55.07 | 58.27 | 64.49 | 55.07 | 58.27 |
| Pancreatic Cancer | 32.71 | 30.02 | 28.27 | 72.90 | 79.32 | 65.38 | 42.06 | 45.33 | 39.38 | 66.36 | 70.18 | 59.38 |
| Breast Cancer | 22.43 | 17.30 | 5.87 | 84.11 | 85.69 | 62.22 | 48.60 | 53.28 | 25.84 | 64.49 | 64.61 | 35.62 |
| Bladder Cancer | 23.36 | 18.29 | 26.20 | 40.19 | 35.79 | 47.76 | 65.42 | 69.58 | 80.83 | 47.66 | 45.92 | 57.91 |
| Chronic Lymph Leukaemia | 21.50 | 16.50 | 27.12 | 49.53 | 45.53 | 64.58 | 45.79 | 41.15 | 58.35 | 49.53 | 44.14 | 63.02 |
| Basal Cell Carcinoma | 69.16 | 55.86 | 89.90 | 11.21 | 7.55 | 70.61 | 14.02 | 10.54 | 73.08 | 22.43 | 14.12 | 75.16 |
| Type 2 Diabetes | 50.47 | 60.44 | 13.62 | 64.49 | 71.37 | 16.29 | 62.62 | 68.79 | 15.58 | 34.58 | 45.33 | 9.90 |
| BMI | 37.38 | 36.78 | 8.83 | 85.98 | 83.30 | 27.56 | 57.01 | 58.85 | 15.69 | 40.19 | 38.77 | 9.50 |
| Melanoma | 28.04 | 14.51 | 76.96 | 21.50 | 9.74 | 73.92 | 32.71 | 22.86 | 79.95 | 32.71 | 23.26 | 80.15 |
| Ischaemic Stroke | 22.43 | 26.24 | 5.59 | 88.79 | 82.90 | 28.67 | 65.42 | 64.02 | 18.41 | 38.32 | 38.17 | 8.67 |
| Glioma | 0.93 | 4.57 | 13.26 | 49.53 | 42.35 | 64.70 | 23.36 | 18.69 | 39.90 | 3.74 | 5.96 | 15.69 |
| Lung Cancer | 7.48 | 13.32 | 10.38 | 14.95 | 25.05 | 21.09 | 11.21 | 22.66 | 18.97 | 23.36 | 37.38 | 36.34 |
| Multiple myeloma | 7.48 | 5.37 | 3.31 | 20.56 | 17.50 | 10.98 | 18.69 | 15.71 | 10.10 | 46.73 | 44.53 | 31.15 |
| Atrial Fibrillation | 24.30 | 16.50 | 5.07 | 23.36 | 15.51 | 4.83 | 13.08 | 8.55 | 2.44 | 9.35 | 7.16 | 2.00 |

**Table 9:** Correlation coefficients between 27 trait percentiles in each distribution within a family memsber.

| *Percentile correlation within Father* | IBS (107) | EUR (503) | ALL (2504) |
| --- | --- | --- | --- |
| IBS (107) | 1 |  |  |
| EUR (503) | 0.9748182 | 1 |  |
| ALL (2504) | 0.801632 | 0.741393034 | 1 |

| *Percentile correlation within Mother* | IBS (107) | EUR (503) | ALL (2504) |
| --- | --- | --- | --- |
| IBS (107) | 1 |  |  |
| EUR (503) | 0.97633965 | 1 |  |
| ALL (2504) | 0.46754615 | 0.439708 | 1 |

| *Percentile correlation within Daughter* | IBS (107) | EUR (503) | ALL (2504) |
| --- | --- | --- | --- |
| IBS (107) | 1 |  |  |
| EUR (503) | 0.9766052 | 1 |  |
| ALL (2504) | 0.6405275 | 0.587166432 | 1 |

| *Percentile correlation within Son* | IBS (107) | EUR (503) | ALL (2504) |
| --- | --- | --- | --- |
| IBS (107) | 1 |  |  |
| EUR (503) | 0.96899774 | 1 |  |
| ALL (2504) | 0.70419702 | 0.658489 | 1 |

## Discussion

Approaches combining the information from large numbers of genomic variants into PRS promise substantial improvement of risk prediction for common diseases and cancer [113]. Implementation of PRS at scale in health services, however, remains a challenge, particularly the translation of PRS into actionable benefits for individuals. Governments in various countries, including in the UK [2], have the ambition to use PRS in healthcare settings, which implies that existing PRS studies do need to be translated into actionable tools for use at the individual level. Such translation requires standardisation so that implementation can be scaled to large numbers of people. In this paper we developed a proof-of-principle implementation of publicly available PRS information, following a systematic curation, deployment and translation of PRS into personalised risk assessments, using a family of four as a test case. A selection of 27 common diseases and cancers (phenotypes) resulted from our curation process, encompassing almost 40 million SNPs. We applied PRS to 1000 Genomes Project (1000G) participants, using the effect weights of over 96 billion risk alleles to construct a background distribution of 27 PRS from which to infer risk percentiles for each of our four family members.

Our curated set of PRS from 27 diverse conditions span autoimmune, cancer, cardiovascular and metabolic diseases. In what follows we discuss our PRS curation, risk percentile generation and interpretation of disease risk assessments as well as opportunities and limitations that such a PRS implementation provides for disease prevention.

### PRS deployment requires considerable curation

Among the many hundreds of PRS we found in online repositories, we note varying study designs, PRS performances, validation cohorts and risk metrics. While this makes it challenging to deploy existing PRS data into a coherent framework for testing of individuals, it also reflects the diverse study designs and analysis methods of the original studies. We developed a set of curation criteria, allowing us to shortlist candidate PRS. Our own judgement was needed in order to evaluate their final inclusion in our analysis. This meant that our selection criteria were not always strictly followed, reducing the potential for standardisation and scalability.

### Percentile PRS calculation lies at the core of risk inference

The concept of putting the individual into the context of a wider population has allowed us to posit a template for turning PRS developed at the population level into a tool which can be applied to individuals. We believe in this approach, largely because the risk metric which results is one of relative risk, consistent with the methodology underpinning the PRS validation process.

By calculating the PRS for each individual within the 1000G, and then placing the family members within the context of that distribution, a robust method of translating population level PRS into relevant individually related scores was arrived at. This is further supported by using whole genome sequencing, and thus avoiding the need for imputation of alleles at any given PRS SNP, which we expect to provide more accurate results. Moreover, from a total of almost 40 million SNPs in 27 phenotypes, 99.97% passed our bioinformatics curation criteria, which allowed us to reliably implement the published PRS in both our tested individuals and background 1000G populations.

In addition, background populations were independent from the cohorts used for training and validation of the PRS. This allowed us to independently test the effect of the choice of background population (IBS, EUR and ALL) for percentile calculation.

### Role of background population in percentile calculation

To ascertain whether the choice of background population had any effect in percentile calculation, we checked how correlated phenotype percentiles are. We found that when considering all phenotypes together, for each family member correlations between IBS and EUR background population results are high. This is perhaps not surprising; the family members are of Iberian Spanish origin, IBS is a subset of the EUR population, and should also be ethnically closer than other groups to the validation populations used by the genetic risk scores themselves and the underlying GWAS studies on which those validations are based.

Perhaps more interesting is what we found when looking at the correlations between ALL, and the other two groups, EUR and IBS. Here we saw markedly lower correlations. On the surface, this could again be expected. ALL contains population groups ethnically very distinct from EUR or IBS, for example East Asians or Africans. However, when we look at the individual phenotypes, a slightly more nuanced picture emerges. The quintile analysis revealed that the results for some phenotypes are consistent between ALL and EUR or IBS (e.g. Coronary Artery disease and Colorectal Cancer) and wholly different in other phenotypes (e.g. Type 2 diabetes and Basal Cell Carcinoma). In fact, it is likely that the moderate correlation we see between ALL and EUR or IBS across all phenotypes is a function of averaging across the selected group of phenotypes, which hides a wide range of differences across populations.

We note that studies with multiple PRS (e.g. Khera et al. (2018), Cancer-PRSweb, etc.) contain phenotypes differing by >20 percentile points for the same individual across population groups, suggesting that the results we are seeing are not due to study design.

In order to explain this consistency between ALL and IBS or EUR for some phenotypes and inconsistency for others, we can hypothesise that for ‘consistent’ quintile PRS percentiles, the frequencies of variants of their PRS SNPs are conserved across the different populations. This may mean that such PRS are more portable than others across different ancestry groups, however we stress that this is speculation, and would require further work. Such work might seek to validate the more ‘consistent’ PRS in non-European population groups. In turn, this validation would require access to (and the existence of) large scale biobank data in such populations, which remains a challenge.

### Translation of percentiles into risk metrics

Our method for translating PRS percentiles into risk metrics relied on their availability in source publications. In order for us to reuse source publication risk metrics they had to be associated to PRS percentile interval thresholds. We found risk metrics to be variably reported, with some studies reporting odds ratios, others hazard ratios and others still no risk metric at all. Furthermore, some studies reported risk over the 80th PRS percentile threshold while others over the 75^th^ or even the top 50^th^ percentile. Still others reported risk of one group relative to a reference group (for instance Mars et al. [21]), rather than relative to the rest (for instance Khera et al. [3]).

We translated genetic risk regardless of thresholds wherever available, but it was not possible to follow a uniform set of rules with which to report risk. For future developments, a standard set of thresholds and risk metrics with which to report genetic risk would be highly desirable for at scale implementation. This would allow for direct comparison between different PRS, creating a common basis for a discussion about disease risk for complex genetic disorders.

Additionally, when considering healthcare interventions, similar phenotype odds ratios with different AUC performances may lead to different levels of confidence. To illustrate this point, we can compare two different, high quality approaches, in Khera et al. [3] and Abraham et al. [108]. The Coronary Artery Disease PRS AUC in Khera et al. has an AUC of 0.81, suggesting that it is able to stratify individuals into different risk bins with a good level of accuracy. With such a robust PRS, certain preventative healthcare interventions for an individual in a high-risk bin might be justified by the PRS alone (for instance lifestyle adjustments). By contrast, Abraham et al.’s Ischaemic Stroke PRS [108] has a C-index of 0.58. With a significantly less robust PRS such as this, it is harder to justify intervening based on the PRS alone, even if the individual in question shows up in a high-risk part of the distribution, as there is less confidence that the risk is correctly attributed to that individual. However, that does not mean that such a PRS is without use. As Abraham et al. (2019) point out, the Ischaemic Stroke PRS with a C-index of 0.58 is still comparable to other common predictors of Ischaemic Stroke, for instance, family history of stroke (C-index of 0.56) Systolic Blood Pressure (C-index of 0.57) or BMI (C-index of 0.57). Therefore, while this PRS is not robust enough to be used on a standalone basis, it nonetheless adds value to the overall assessment of risk of Ischaemic Stroke, and when combined with other risk factors including hypertension, raises the C-index to 0.635 [108].

### Limitations of this implementation

First and foremost, our analysis is limited by the size of our use case, the family of four. As a result, we seek only to offer a proof of concept, and some insights which we believe are generalisable to implementations at a larger scale. A greater number of subjects would be needed in order to validate the specific results presented here.

The second important limitation is population constraints. Our percentile risk calculation has only been performed for an Iberian family. A subsequent analysis could consider individuals from different ancestry backgrounds against different population cohorts. Further, our results may have been affected by the way that the 1000G population groups are constructed. The same Iberian Spanish participants of the 1000G are included in the European subset population, and in turn, all Europeans are included in the total population of the 1000G (ALL). We also note that the Iberian population is of small size (n=107) in the 1000G, reducing the statistical significance of results using that population as background distribution. We are also conscious that all PRS used here were themselves derived from and validated in Northern European populations (White British or Finish), which may also contribute inaccuracy to our risk analysis in the IBS subpopulation.

There are also a number of limitations imposed by bioinformatics constraints. We require the overwhelming majority of PRS SNPs to be present in the 1000G population, which may rule out some high quality PRS. Furthermore, we were only able to select PRS whose number of SNPs not present in the 1000G dataset was smaller than 5% of the total. Missing SNPs in the PRS calculation will have a greater impact for phenotypes where the PRS had few SNPs (e.g., Basal Cell Carcinoma is the phenotype that proportionally misses the greatest proportion of SNPs, 1 out of 23; 4.35%), in contrast to those that included all SNPs genome wide (e.g., Type 2 Diabetes ∼7 million SNPs). We nevertheless believe that the expected impact is not significant, since for the great majority of PRS we have used, considerably less than 5% of the SNPs were missed or none at all (see Table 4, ‘% Missing SNPs’). Another weakness of our current methodology is that we have excluded PRS that contained SNPs in the X and Y chromosomes, resulting in more missing SNPs in certain phenotypes, and so causing us to exclude them (e.g. Testosterone Levels.

## Conclusion

We have presented a comprehensive set of 27 curated PRS encompassing autoimmune, metabolic, cancer and cardiovascular diseases. We offer a proof-of-principle approach for an implementation of individualised PRS analysis, with a test case of a family of four using background distributions from 1000G. These 1000G populations allow us to calculate PRS and extrapolate them into relative risk for individuals using as input whole genome variant data. Calculated risk percentiles from PRS allow us to infer relative risks for any of the diseases analysed here. We show how current lack of standards for risk reporting challenges our ability to implement PRS more straightforwardly. It is also noted that different disease risks cannot be uniformly interpreted as their differences in study design, performances and risk reporting are not standardised. We further explore the effect of background population on an individual PRS percentile by comparing how different 1000G populations affect resulting PRS percentile calculations. All in all, this work offers insight into how PRS can be translated into relative risks for individuals, and therefore showcases their potential for their deployment in a preventative healthcare setting.

## Data Availability

The sources of PRS used in this study are included in Table 3 and Table 4. The family genome variation data for this manuscript is not publicly available because they were not consented for open access. Request to access the family genome variation data should be directed to Manuel Corpas.

## Author contributions

MC and EL conceived the experiments and performed the analysis. MC wrote the paper with contributions from all authors. All authors read and approved the paper.

